# Decreased plasma levels of the survival factor renalase are associated with worse outcomes in COVID-19

**DOI:** 10.1101/2020.06.02.20120865

**Authors:** Melinda Wang, Xiaojia Guo, Hyung J Chun, Alfred Ian Lee, Charles Cha, Fred Gorelick, Gary V Desir

## Abstract

**Introduction:** Renalase (RNLS), a novel secreted plasma flavoprotein, has anti-inflammatory effects in a variety of disease processes. Severe COVID-19 disease is associated with disordered inflammatory responses. We hypothesized that reduced plasma RNLS levels could be a marker of COVID-19 disease severity.

**Methods:** Plasma was collected from 51 hospitalized COVID-19 patients and 15 uninfected non-hospitalized controls. Plasma RNLS and cytokine levels were measured and sociodemographic and clinical data were collected from chart review. Data were analyzed using nonparametric analyses and Kaplan Meir curve log rank analysis.

**Results:** Plasma RNLs levels were negatively correlated with inflammatory markers, including IL-1β, IL-6, and TNFα (p = 0.04, p = 0.03, p = 0.01, respectively). Patients with COVID-19 disease had lower levels of RNLS than controls. Lower levels of RNLS were associated with more severe disease among COVID-19 patients. Low RNLS was also associated with worse survival among COVID-19 patients (HR = 4.54; 95% CI: 1.06-19.43; p = 0.005).

**Conclusion:** Low plasma RNLS levels are associated with severe COVID-19 disease and may be a useful additional biomarker when identifying patients with severe COVID-19 disease. Given RNLS’ antiinflammatory properties and negative correlation with inflammatory markers, these findings also suggest evidence of a potential pathophysiological mechanism for severe COVID-19 disease.

## Introduction

To date, approximately 6 million global cases of COVID-19 have caused ~375,000 death.^1^ The disease ranges from asymptomatic to severe cases characterized by extensive lung injury and an intense systemic hyperinflammation response.^2^ Validated markers of disease progression could affect management and therapy.

Renalase (RNLS), a protein highly expressed in kidney and secreted into blood, displays potent prosurvival and anti-inflammatory effects. In acute kidney injury,^3^ and acute pancreatitis,^4^ reduced blood RNLS leads to increased organ injury and inflammation. Cellular injury, disordered inflammatory responses, increased inflammasome-driven pathways and reduced T-cell function together are central to the pathogenesis of COVID-19.^5^ Given RNLS’s ability to reduce tissue injury and inflammation, we hypothesized that its plasma levels could be a marker of COVID-19 disease severity.

## Methods

Plasma was collected from 51 adult COVID-19 patients hospitalized at Yale New Haven Hospital from March 1 to April 23, 2020, and 15 non-hospitalized, uninfected controls. Thirty of 39 (76.9%) ICU patients were mechanically ventilated at enrollment. All 12 non-ICU patients were on ≤ 3 L supplemental oxygen. The protocol was approved by the local institutional review board (HIC 2000027792).

Plasma RNLS levels were assayed as the denaturation (acid)-sensitive pool by ELISA as described.^6^ Patients were categorized as having low or high RNLS levels (defined as values within or above the bottom quartile respectively). Inflammatory markers, including IFNγ, IL-1β, IL-6, and TNFα were measured using the same blood sample using V-Plex Proinflammatory Panel 1 kits (Meso Scale Diagnostics, LLC, Rockville, MD), according to manufacturer’s instruction. All other laboratory tests were measured from clinical samples on the same day as plasma RNLS lab draw. Sociodemographic data were derived from medical records. Analysis was conducted using nonparametric statistical methodologies (SPSS Version 24, IBM Statistics; Armonk, NY). Kaplan-Meier curve survival analysis was completed using log rank analysis. P-value ≤ 0.05 was considered statistically significant.

## Results

In the COVID-19 cohort (n=51), RNLS levels ranged from 1,964 ng/mL to 22,675 ng/mL with a median of 7,144 ng/mL; the bottom quartile cut-off was 5,522 ng/mL. We compared subjects with high to those with low RNLS levels, and found no significant differences in age, gender, body mass index, race, comorbidities, use of steroids or tocilizumab, eGFR, and D-dimers levels **(Table 1)**. Patients with low RNLS levels were more likely to be treated with remdesivir than patients with high RNLS levels. All patients treated with remdesivir (n = 6) were ICU patients. Plasma RNLS levels were negatively correlated with IL-lβ (r = -0.26, p = 0.04), IL-6 (r = -0.27, p = 0.03), and TNFα (r = -0.31, p = 0.01) but not with IFNγ (r = -0.03, p = 0.80) (**Figure 1**). Patients with COVID-19 had significantly lower plasma RNLS levels than controls **(Figure 2a)**. Of 39 subjects requiring ICU management, 12 (31%) had plasma RNLS levels below the minimum value of non-ICU patients **(Figure 2b)**. COVID-19 subjects with low RNLS levels were significantly more likely to require mechanical ventilation compared to those with high RNLS levels (84.6% vs. 50.0%, p = 0.048). Low RNLS levels were associated with worse overall survival (HR = 4.54; 95% Cl: 1.06-19.43; p = 0.005; **Figure 3)**. Excess mortality remained significant in COVID-19 subjects when low RNLS was defined as the bottom tertile (tertile 1 vs. 2,3: p = 0.03; 1 vs. 2: p = 0.04; 1 vs. 3: p = 0.05).

**Table 1.**

| **Factor** | **Low Bound RNLS**<br>**(n= 13)** | **High Bound RNLS**<br>**(n= 38)** | **p-value** |
| --- | --- | --- | --- |
| Age | 57.82 (35.92-72.25) | 63.55 (33.97-86.31) | 0.489 |
| Gender |  |  | 1.000 |
| Male | 9 (69.2%) | 25 (65.8%) |  |
| Female | 4 (30.8%) | 13 (34.2%) |  |
| BMI | 35.64 (20.69-38.74) | 34.00 (20.82-44.98) | 0.820 |
| Race |  |  | 0.450 |
| White | 5 (38.5%) | 23 (60.5%) |  |
| Black | 5 (38.5%) | 8 (21.1%) |  |
| Asian | 0 (0.0%) | 1 (2.6%) |  |
| Other | 3 (23.1%) | 6 (15.8%) |  |
| Hypertension | 6 (53.8%) | 20 (52.6%) | 1.000 |
| CHD | 4 (30.8%) | 8 (21.1%) | 0.474 |
| CKD | 2 (15.4%) | 4 (10.5%) | 0.638 |
| Chronic lung disease | 1 (7.7%) | 5 (13.2%) | 1.000 |
| Immunocompromised | 4 (30.8%) | 5 (13.2%) | 0.208 |
| Cancer | 3 (23.1%) | 4 (10.5%) | 0.352 |
| T2DM | 4 (30.8%) | 12 (31.6%) | 1.000 |
| Hydroxychloroquine | 13 (100.0%) | 33 (86.8%) | 0.311 |
| Remdesivir | 4 (30.8%) | 2 (5.3%) | **0.031** |
| Steroids | 8 (61.5%) | 20 (52.6%) | 0.749 |
| Tocilizumab | 12 (92.3%) | 31 (81.6%) | 0.662 |
| eGFR | 60 (10-60) | 60 (11-60) | 0.545 |
| D-Dimer | 2.64 (0.17-18.61) | 3.95 (0.19-33.70) | 0.426 |

**Figure 1.**
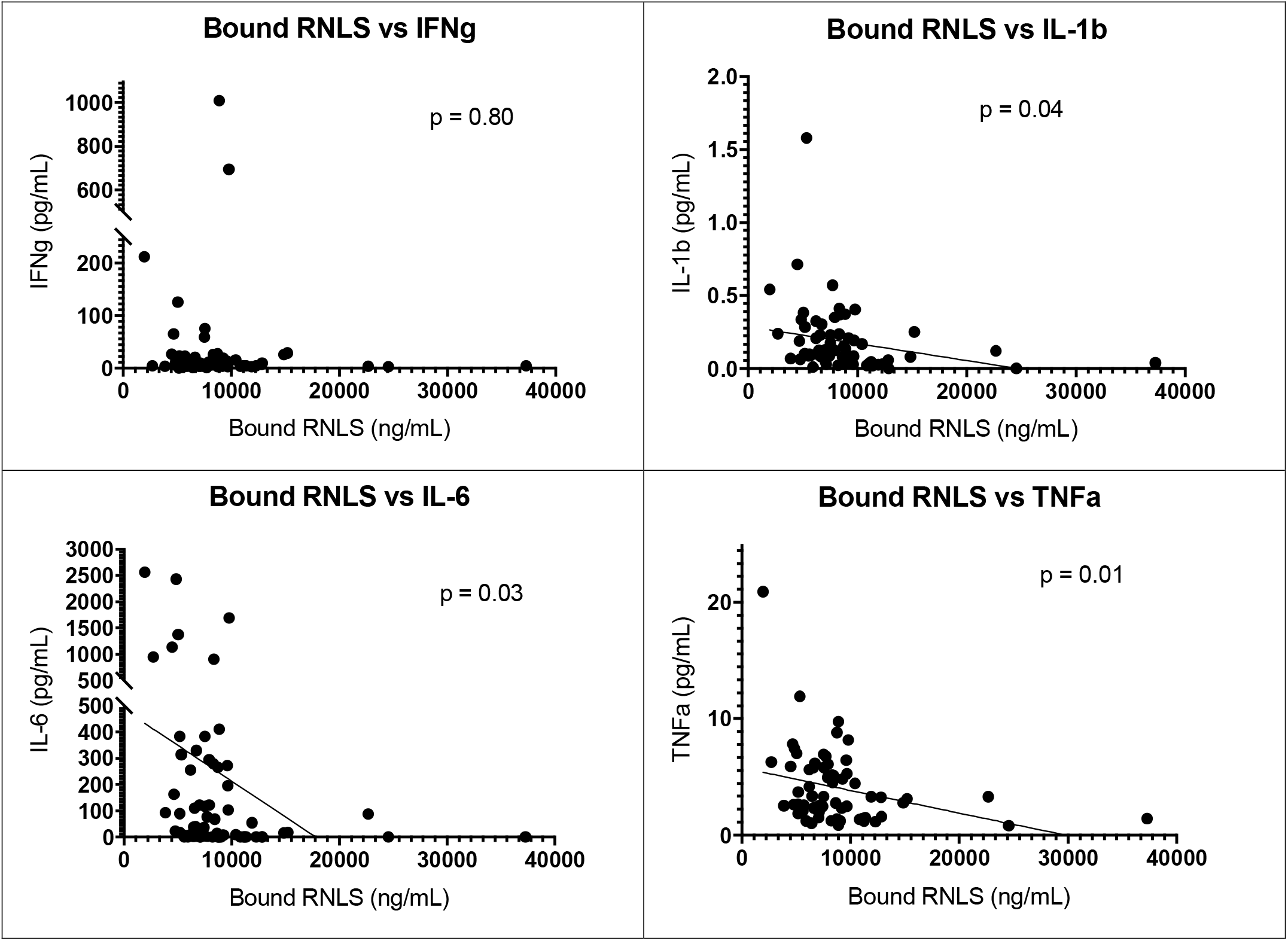
Plasma RNLS levels are negatively correlated with inflammatory markers.

**Figure 2.**
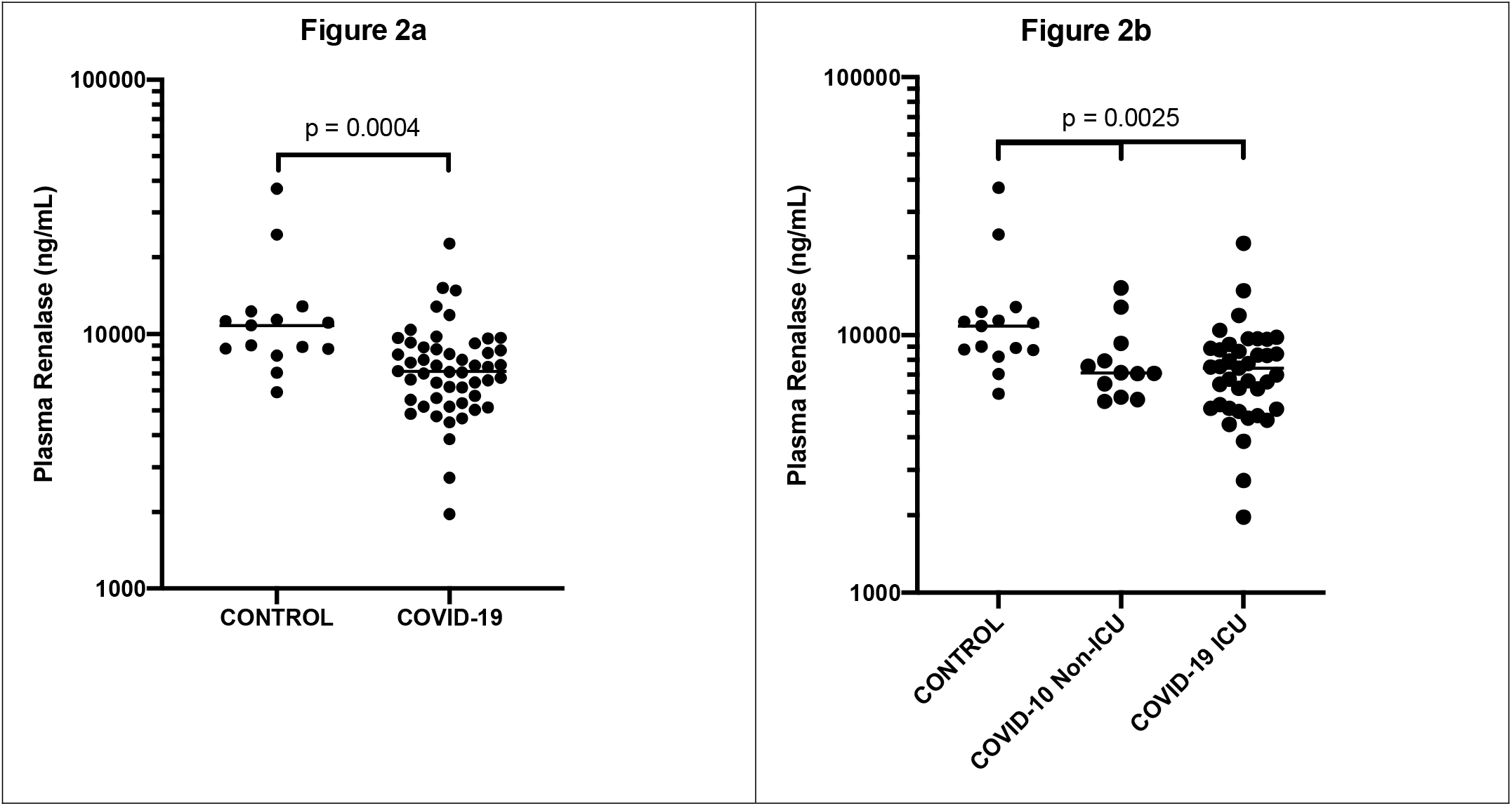
Plasma RNLS Levels are lower in hospitalized COVID19 patients with and without ICU care than healthy controls. **A-** COVID19 negative controls: n=15; COVID19 (+) n=51; **B-** COVID19 hospitalized without ICU care: n=12; with ICU care: n=39

**Figure 3.**
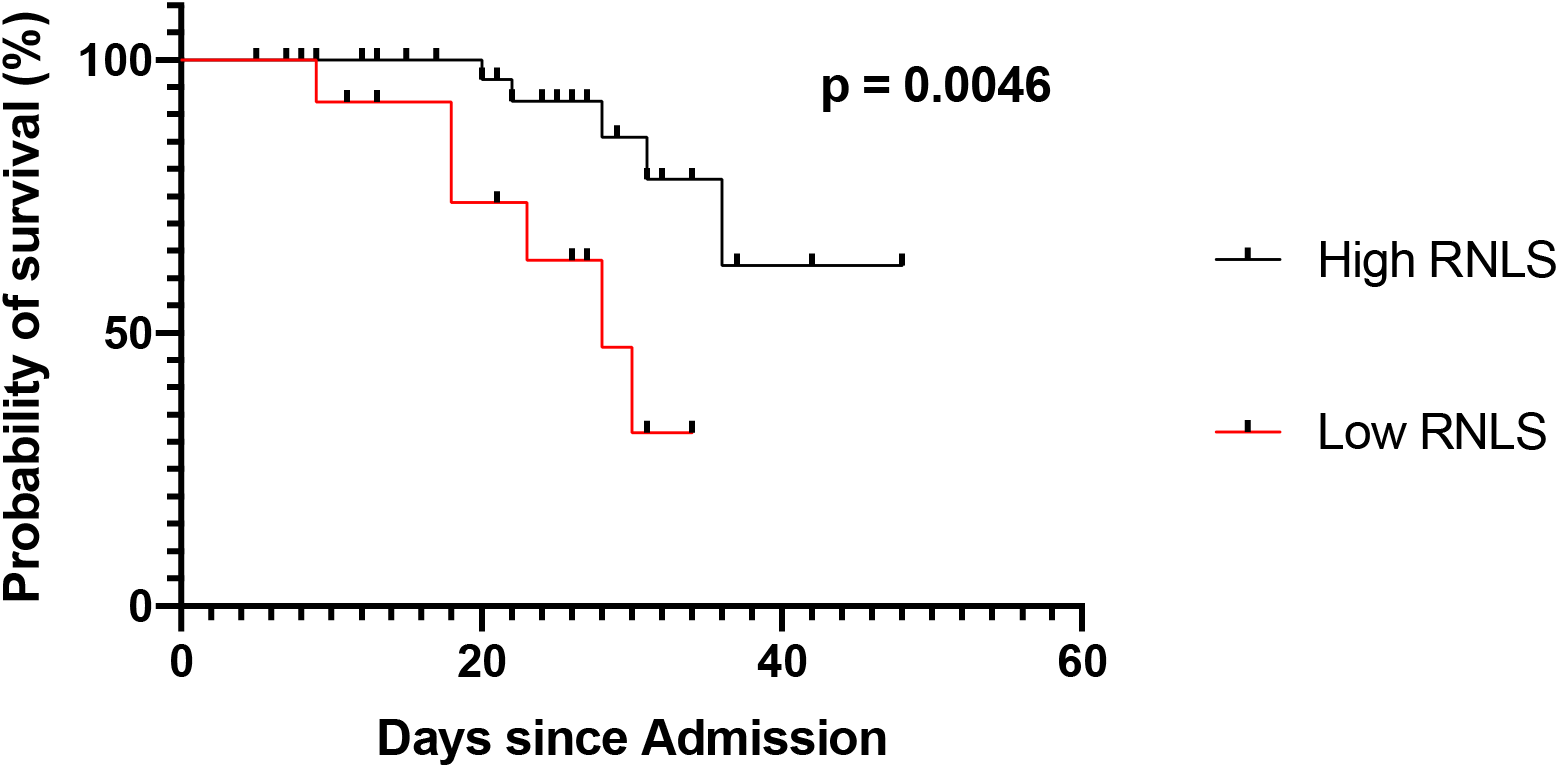
High plasma RNLS levels confer a survival advantage in severe COVID19. Kaplan-Meir Survival Curve in COVID19 patients with high (n=37) vs. low plasma (n=14) RNLS levels.

## Conclusion

In hospitalized COVID-19 patients, low plasma RNLS levels were associated with a greater likelihood of requiring mechanical ventilation and worse overall survival. Although remdesivir treatment was associated with low plasma RNLS levels, all patients treated with remdesivir were admitted to the ICU, suggesting remdesivir use in our cohort likely correlated with more severe COVID19 disease. Plasma RNLS levels were negatively correlated with common inflammatory markers, including IL-1β, IL-6, and TNFα. This finding corresponds with studies showing the anti-inflammatory properties of RNLS.^3,4^ Low RNLS levels may therefore be associated with increased inflammation and more severe COVID19 disease. To our knowledge, this is the first study to report an association between plasma RNLS levels and COVID-19 severity. Limitations of our study include a small sample size, recruitment from a single institution, and potential bias in patient selection methodology. Nevertheless, its results suggest that plasma RNLS may be useful as an additional biomarker to identify patients with more severe disease and alludes to a potential pathophysiological mechanism associated with inflammation in COVID19 disease.

In summary, these results are consonant with published studies that support the concept that sufficient plasma levels of renalase are critical for maintenance of biologic integrity in a broad range of diseases. Low plasma renalase may be a cause and/or marker for the systemic biologic perturbations that lead to a poor outcome in COVID-19. Accordingly, the potential therapeutic value of RNLS agonist administration in patients with severe COVID-19 should be examined.

## Authors Contributions

**Concept and design:** Desir and Gorelick

**Acquisition, analysis:** Lee, Chun, Wang, Guo, Cha

**Interpretation:** Gorelick, Desir, Cha

**Drafting of manuscript:** Wang, Gorelick, Desir

Review of manuscript: Guo, Lee, Chun, Cha

**Drs. Desir and Gorelick:** full access to the study data

## Data Availability

The data referred to in this manuscript are not currently available.

## Acknowledgements

Our sincere thanks to Drs. Aldo Peixoto and Robert Soufer for their advice and careful review of the manuscript.

